# Mortality reduction in ICU-admitted COVID-19 patients in Suriname after treatment with convalescent plasma acquired via gravity filtration

**DOI:** 10.1101/2021.04.14.21255104

**Authors:** R. Bihariesingh-Sanchit, R. Bansie, J. Fröberg, N. Ramdhani, R. Mangroo, D. Bustamente, E. Diaz, I. Thakoer, S. Vreden, Z. Choudhry, W. Jansen Klomp, D.A. Diavatopoulos, A. Nierich

## Abstract

**Background:** Although convalescent plasma (CP) treatment is a potential therapeutic option for patients with COVID-19, there is a paucity of data from studies in low-resource settings. The efficacy and safety of CP therapy in intensive care unit (ICU) patients with COVID-19 in Suriname was evaluated. A novel gravity-based filtration method was used to obtain CP. The design was an open-label, multi-centre, non-randomized prospective clinical trial performed in two hospitals in Suriname, from June 2020, to December 2020. A pre-planned interim analysis is reported in 78 PCR-confirmed COVID-19 patients admitted to the ICU with severe or life-threatening symptoms. CP in combination with standard treatment (n = 28) was compared to standard treatment alone (control) (n = 50), stratified by disease severity. The primary endpoint was 28-day ICU mortality. Secondary (exploratory) endpoints were changes two days after treatment initiation in pulmonary oxygen exchange capacity (PF ratio) and chest x-ray (CXR) score.

**Findings:** The median age of patients was 52 years with 43 [55.1%] being male. Twenty-eight day mortality occurred in 18% (5/28) of the CP group vs 36% (18/50) of the control group. Survival probability was significantly higher in the CP group compared to the control group with standard care (P=0.027). When stratifying into disease severity, the survival probability was the lowest for the control group with life-threatening disease (P=0.0051). CP treatment of severe COVID-19 resulted in a higher probability of survival, with a hazard ratio (HR) of 0.22 (95% CI, 0.074-0.067), correcting for age, the presence of diabetes and COVID-19 severity. Age significantly increased the mortality risk (HR, 1.08 [95%CI, 1.022-1.14]; P = 0.006). In the severe group, CP resulted in an improved CXR score (P =0.0013) and increase in PF ratio (P = 0.011) as compared to standard therapy. The gravity-based plasmapheresis method used for CP production was well-tolerated and no adverse events were observed in the donors.

**Interpretation:** Among patients with severe or life-threatening COVID-19, CP therapy in combination with standard treatment resulted in 78% reduction of 28-day ICU mortality (HR = 0.22) compared to standard treatment alone. Both CXR-score and PF ratio changes represent indicators of treatment effect of CP after two days and can easily be implemented in low-resource settings. The novel CP production method was effective and represents a practical solution for low- and middle income countries (LMICs) to produce CP locally. Although interpretation is limited by the non-randomized design of the trial, these results offer a potential route for broader implementation of CP treatment in LMICs.

**Funding:** This study was funded by the Academic Hospital Paramaribo, without additional third-party funding.

## Introduction

The current COVID-19 pandemic has caused a global socio-economical and health care crisis with more than 116,000,000 cases worldwide and over 2,600,000 recorded deaths since the discovery of the Severe Acute Respiratory Syndrome Coronavirus 2 (SARS-CoV-2) in December 2019. The clinical deterioration resulting in tachypnea and dyspnea in approximately 20% of patients is the major complication experienced by patients and physicians today. Treatment of COVID-19 still primarily relies on adequate supportive care, with dexamethasone and remdesivir included in current treatment protocols^1–3^.

The use of convalescent plasma (CP) from patients recovered from COVID-19 as a means of passive antibody therapy has been explored in several clinical trials and has received emergency use authorization in the USA. Although further clinical trials remain necessary to further assess the possible benefit of CP, the FDA has allowed use of this treatment outside of clinical research (FDA August 2020), representing the only investigational product thus far with such a status. Already early during the pandemic, a small number of studies reported promising results on the efficacy of CP treatment in patients with COVID-19 in different clinical settings^4,5^. Despite multiple studies, including retrospective cohort studies, reporting on significant benefits of CP treatment, two randomized controlled trials at the beginning of 2020 showed no overall clinical benefit ^6,7^, with both studies being terminated prematurely. With a population just over 500,000 inhabitants and neighboring Brazil where high COVID-19 disease incidence was reported, Suriname was confronted with a second COVID-19 wave at the end of May 2020. Given the limited treatment options, we initiated a clinical trial to evaluate clinical efficacy of CP treatment in patients with severe or life-threatening COVID-19 in Suriname (SuriCovid trial).

Prior to initiating such a study in the low-resource setting of Suriname, significant logistical challenges had to be to overcome, including the lack of conventional plasmapheresis machines and lack of a local immune assay to quantify virus-specific and virus-neutralizing antibodies (Abs). Furthermore, the optimal volume of CP for infusion as well as safety of blooddonation for a CP donor were unknown. In this study, we made use of a gravity-driven blood filter for CP production, called the HemoClear device^8^. To be able to identify short-term treatment effects in the absence of extensive laboratory diagnostic capacity, changes in oxygen balance and chest x-ray score (CXR) were assessed 2 days after CP therapy.

Here, we present the pre-planned interim analysis on Intensive Care Unit (ICU) outcome in COVID-19 patients in Suriname after CP treatment.

## Methods

This work is reported in adherence to the preferred STrengthening the Reporting of OBservational studies in Epidemiology (STROBE) guidelines. This open-label, non-randomized prospective clinical trial was performed at the Intensive Care Unit of Academic Hospital Paramaribo and the Wanica Regional Hospital, Suriname, from June 2020 to December 2020. Ethical approval was granted by the Suriname Ministry of Health’s Ethics Review Board (registration number:IGAP02-482020; ISRCTN18304314) and was performed in accordance with the declaration of Helsinki. Seventy-eight patients were included.

### Study flow chart

After referral to the ICU, the patients were treated by supported therapy such as additional respiratory and circulatory support. After being found elegible for additional CP treatment, a selection to either standard treatment including dexamethasone or standard treatment added with CP was started. The study flow chart in Figure 1 illustrates the study enrolment and design.

**Figure 1.**
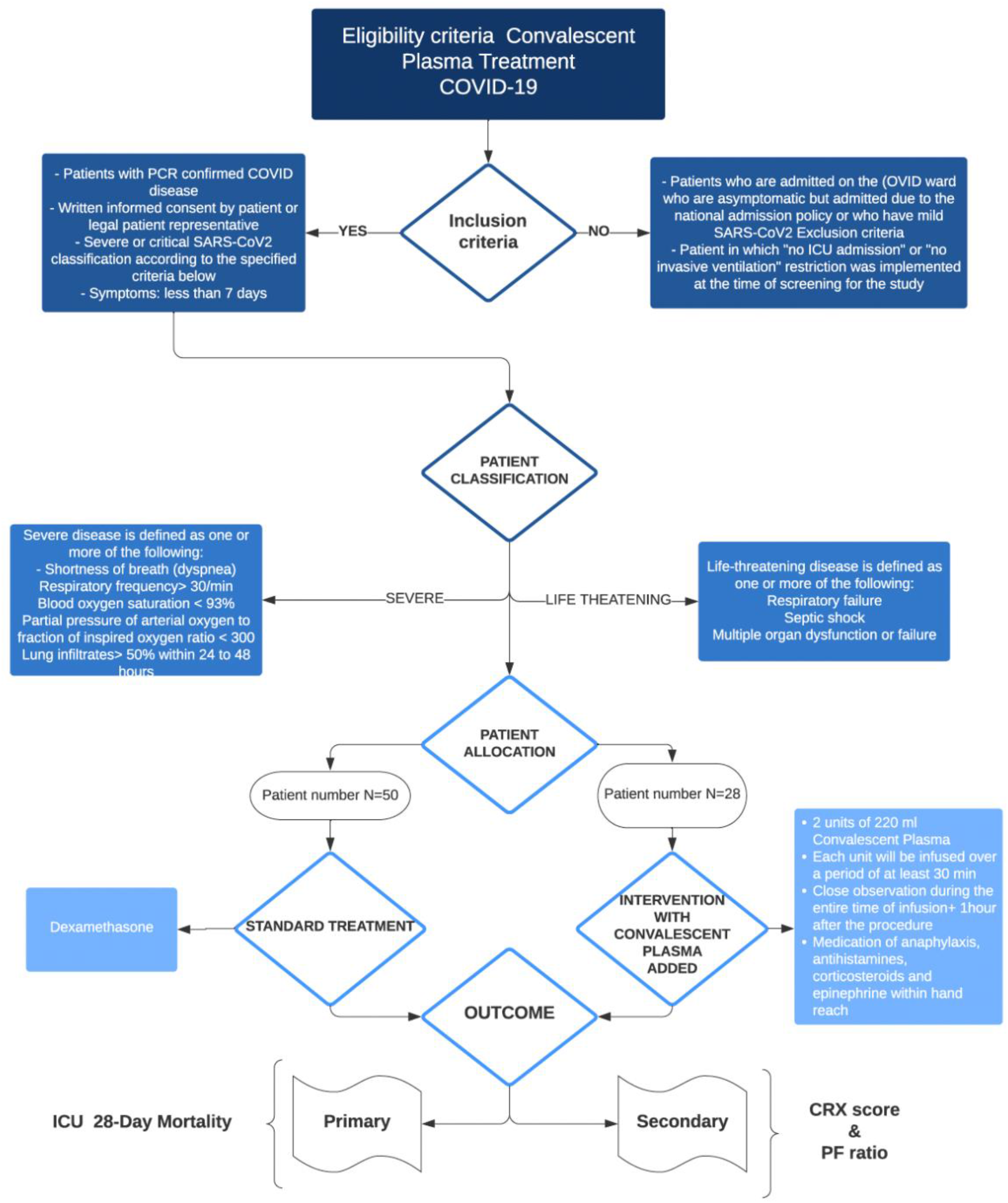
Patient selection Flowchart with study enrolment and design.

### Patients

Adult patients (>18 years) with severe or life-threatening COVID-19 were enrolled in the trial. The eligibility criteria included written informed consent given by the patient or next-of-kin, a PCR confirmed diagnosis of COVID-19, and admittance to the ICU due to progressive respiratory failure ranging between severe and life threatening ARDS based on the Berlin classification^9^. For the interventional CP group, all patients admitted at the ICU who met the inclusion criteria were approached and 28 patients who consented to CP infusion and standard therapy were recruited to this arm. For the control group with standard therapy, patients were included who did not consent to CP infusion or where there was no CP available. To account for major confounding factors, the following variables were used: age, gender, comorbidities including history of diabetes mellitus (DM), hypertension (HTN), drugs, symptoms and signs. In addition, both the controls and treatment groups received the same standard concurrent treatment, which included administration of oral or intravenous dexamethasone at a dose of 6 mg once daily, for up to 10 days. Eligible patients with severe and life threatening COVID-19 were infused with two units of 220 ml CP once with the donated CP. For plasma selection, ABO compatibility was considered, regardless of Rhesus factor status. CP recipients were monitored for serious adverse effects (SAEs) of CP transfusion, including anaphylaxis.

### Data Collection

Data collection forms were developed to collect the data of all participants and included the clinical, radiological and laboratory information that was retrieved from the hospital electronic/paper records system. The data were cross-checked to ensure the minimization of data entry errors by two investigators.

Chest X-rays were obtained at admission on the ICU and after two days of treatment. Chest X-rays were evaluated using the CXR scoring system, which was specifically designed for semi-quantitative assessment of lung involvement in COVID-19^10,11^. In the CXR scoring system, six different lung zones are scored. Each lung zone was scored between 0 and 3, leading to an overall “CXR-SCORE”, ranging from 0 to 18:

- Score 0 no lung abnormalities
- Score 1 interstitial infiltrates
- Score 2 interstitial and alveolar infiltrates (interstitial predominance)
- Score 3 interstitial and alveolar infiltrates (alveolar predominance).

Chest x-rays were examined immediately prior to (day 0) and two days after (day 2) CP administration, or on day 0 and day 2 following inclusion in de the control group by two independent radiologists blinded to group assignment. CXR scoring was based on radiological improvement observed after treatment with convalescent plasma^4,10^.

Ventilation parameters, including pH, pCO2, pO2, FiO2, PaO2 / FiO2, were assessed on day 0 and day 2 for both groups, were used to measure oxygen balance changes as the calculated PaO2/FiO2 (PF) ratio. This is the ratio of arterial oxygen partial pressure (PaO2 in mmHg) to fractional inspired oxygen and is related to severity of disease and outcome by calculation the Acute Respiratory Distress Syndrome scale^9,12^.

### Convalescent plasma donor recruitment

CP donors were recruited from hospitalized adult (≥18y) patients who had recovered from a PCR-confirmed SARS-CoV2 infection at the time of plasma donation. Eligibility criteria for donating CP were a positive response measured as optical densitometry (OD) levels of the anti-SARS-CoV-2 total IgG antibody Wantai test^13^, being at least 14 days asymptomatic following resolution of COVID-19 and having two negative PCR-tests from nasopharyngeal swabs. Prior to CP collection by the critical care team, written informed consent was obtained from all donors. Following CP donation, all CP samples were screened using the standard clinical and laboratory protocol used by the National Blood Bank of the Suriname Red Cross. This includes screening for antibodies against potential transfusion transmitted infectious diseases, i.e. HIV1/2, HBsAg, Hepatitis C virus, syphilis RPR, HTLV1/2 and *Trypanosomi cruzi*. Upon donation, 500ml of CP was obtained from each donor.

### Plasma Preparation Procedure and Quality Control

Convalescent plasma was obtained via plasmapheresis using HemoClear blood filters (HemoClear BV, Zwolle, The Netherlands)^14^. In this method, whole blood is separated via gravity-based sterile filtering through a multi-layered cross-flow membrane module^15^. This offers a high yield of CP without loss of RBCs and was the only available plasmapheresis method. The steps for plasma preparation are illustrated in Figure 2 (and Video S1).

**Figure 2.**
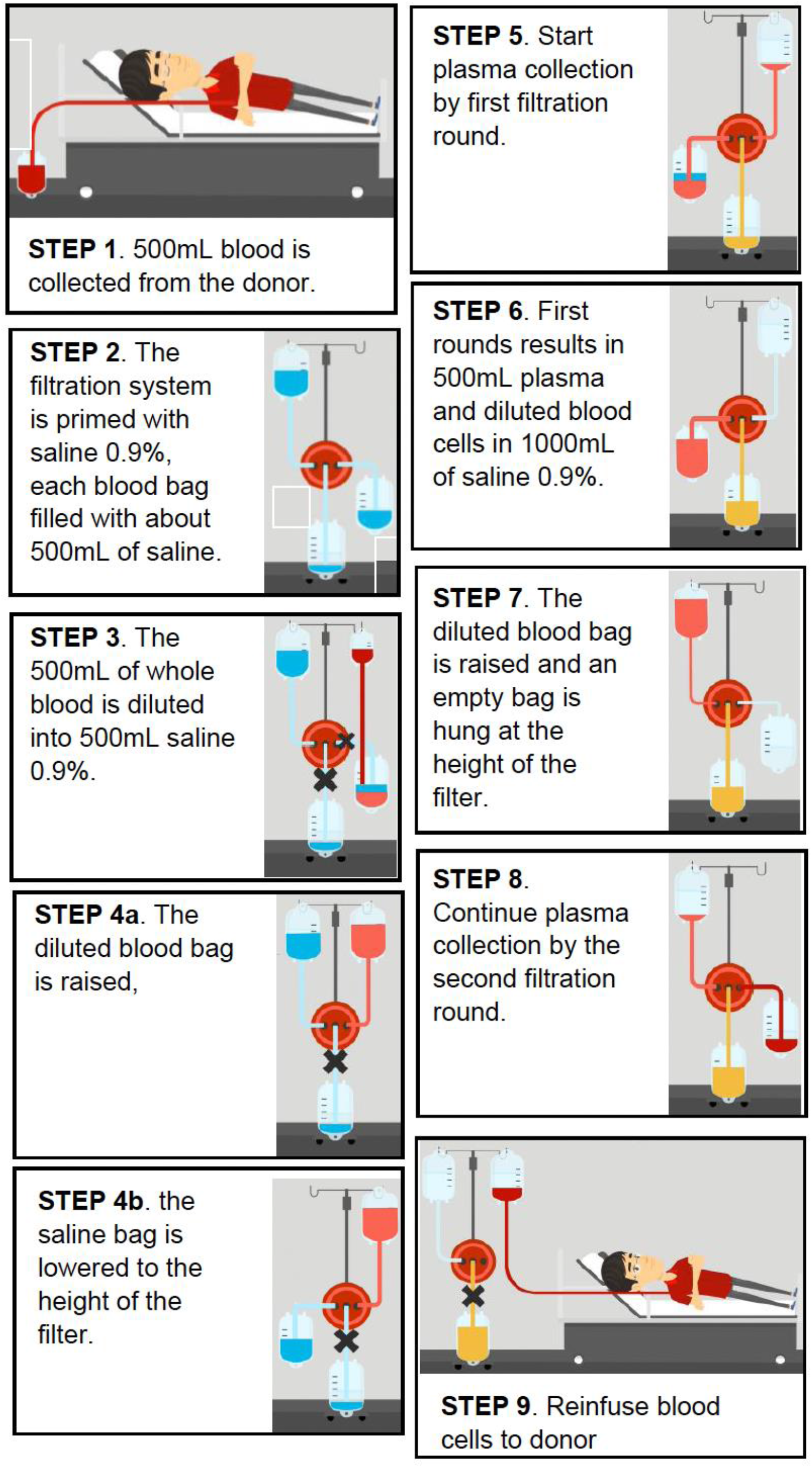
Convalescent plasma collection method by means of the HemoClear system.

By design the filtration process is maintained as a closed-loop system to prevent potential contamination. The diluted blood was passed through the filter during two consecutive filtration rounds. Plasma was collected one meter below the device level in the satellite plasma bag. The blood cell components, diluted with saline 0.9%, were re-infused in the donor. This procedure of blood donation and CP acquisition can then be repeated within the same donor either immediately or on another day. There was no pooled storage after donation. After harvesting, convalescent plasma was sent to the National Blood Bank of the Suriname Red Cross for processing and storage. Plasma was stored at −25C according to the guidelines for plasma storage^16^.

### Statistical Analysis

Unless stated otherwise, all analyses are performed on the complete dataset. General descriptive characteristics were assessed using SPSS for Windows (version 16.0; SPSS). Differences between the two treatment groups were analysed with chi-square or the Fisher exact test where suitable for categorical variables, and t-tests for continuous variables. The differences in outcome measures, the survival analyses and the time to ICU dismissal were performed in R-studio (RStudio Desktop 1.4.1106 open source, Boston, USA). CXR score and PF ratio were summarized by presenting the median and spread in a boxplot. Differences within the therapy group were analysed with a paired t-test, differences between the two groups were analysed with an unpaired t-test. The mortality risk was assessed with the Kaplan Meier method, and hazard ratios were calculated with a Cox proportional hazards model. A P value of 0.05 was considered to represent significant difference. Statistical parameters are reported directly in the figures and figure legends.

## Results

### Patients

A total of 78 patients were enrolled in the trial of which 28 (35.9%) received CP The mean age of the patients was 53.0±14.3 years and 43 patients (55%) were male. There were no significant differences in baseline characteristics between the CP intervention group and the control group. The demographic, clinical factors and laboratory measurements between the intervention (convalescent plasma) group and the control (dexamethasone) group are described in table 1 and 2. Treatment allocation to the intervention group was biased due to limited availability of donors; only part of ICU doctors were organising donations and informed consents of the donors and patients were sceptic about this treatment option based also on religious prohibitive rules. However there was a tendency towards more life-threatening patients allocated to the intervention group than in the standard treatment group (CP 42.9% vs 34%). After initial ICU admission, all patients started with standard treatment.

**Table 1.** Comparison of demographic and clinical factors between the intervention (convalescent plasma) group and the control (dexamethasone) group.

| Variable | Control group<br>(N = 50) | Convalescent<br>plasma<br>(N = 28) | P value |
| --- | --- | --- | --- |
| **Demographic characteristics** |  |  |  |
| Age in years mean (sd) | 52.5 (14.5) | 53.8 (14.3) | 0.71 |
| Male sex n(%) | 30 (60.0) | 13 (48.1) | 0.32 |
| Body mass index, median [IQR] | 30 [28 – 32] | 32 [29 – 35] | 0.19 |
| **Comorbidities n(%)** |  |  |  |
| Diabetes Mellitus | 15 (30.6) | 12 (44.4) | 0.23 |
| Hypertension | 23 (47.9) | 15 (55.6) | 0.53 |
| Stroke | 1 (2.1) | 1 (3.7) | 1.00 |
| Renal disease | 6 (12.5) | 6 (22.2) | 0.33 |
| Ischemic heart disease | 3 (6.4) | 4 (14.8) | 0.25 |
| **Drug usage n(%)** |  |  |  |
| Oral diabetics | 17(35.4) | 10 (38.5) | 0.80 |
| Insulin | 3 (6.4) | 3 (11.1) | 0.66 |
| ACE-inhibitors | 8 (17.0) | 7 (25.9) | 0.36 |
| Angiotensin receptor blockers | 3 (6.4) | 1 (3.7) | 0.62 |
| **Presence of symptoms n(%)** |  |  |  |
| Fever | 39 (78.0) | 22 (81.5) | 0.72 |
| Dyspnea | 44 (88.0) | 24( 88.9) | 1.00 |
| Cough | 25 (50.0) | 18( 69.2) | 0.11 |
| Sputum | 4 (8.2) | 2 (7.4) | 0.91 |
| Loss of taste | 7 (14.3) | 6 (22.2) | 0.53 |
| Anosmia | 7 (14.3) | 5 (18.5) | 0.75 |
| Diarrhea | 10 (20.4) | 3 (11.1) | 0.36 |
| Vomiting | 4 (8.3) | 2 (7.4) | 1.00 |
| Acute renal failure | 3 (6.3) | 6 (23.1) | 0.06 |
| **Covid Classification n(%)** |  |  | 0.44 |
| Severe | 33 (66.0) | 16 (57.1) |  |
| Life-threatening | 17 (34.0) | 12 (42.9) |  |

Abbreviations: ACE: angiotensine Converting Enzyme, IQR: interquartile range, sd: Standard deviation

**Table 2.**
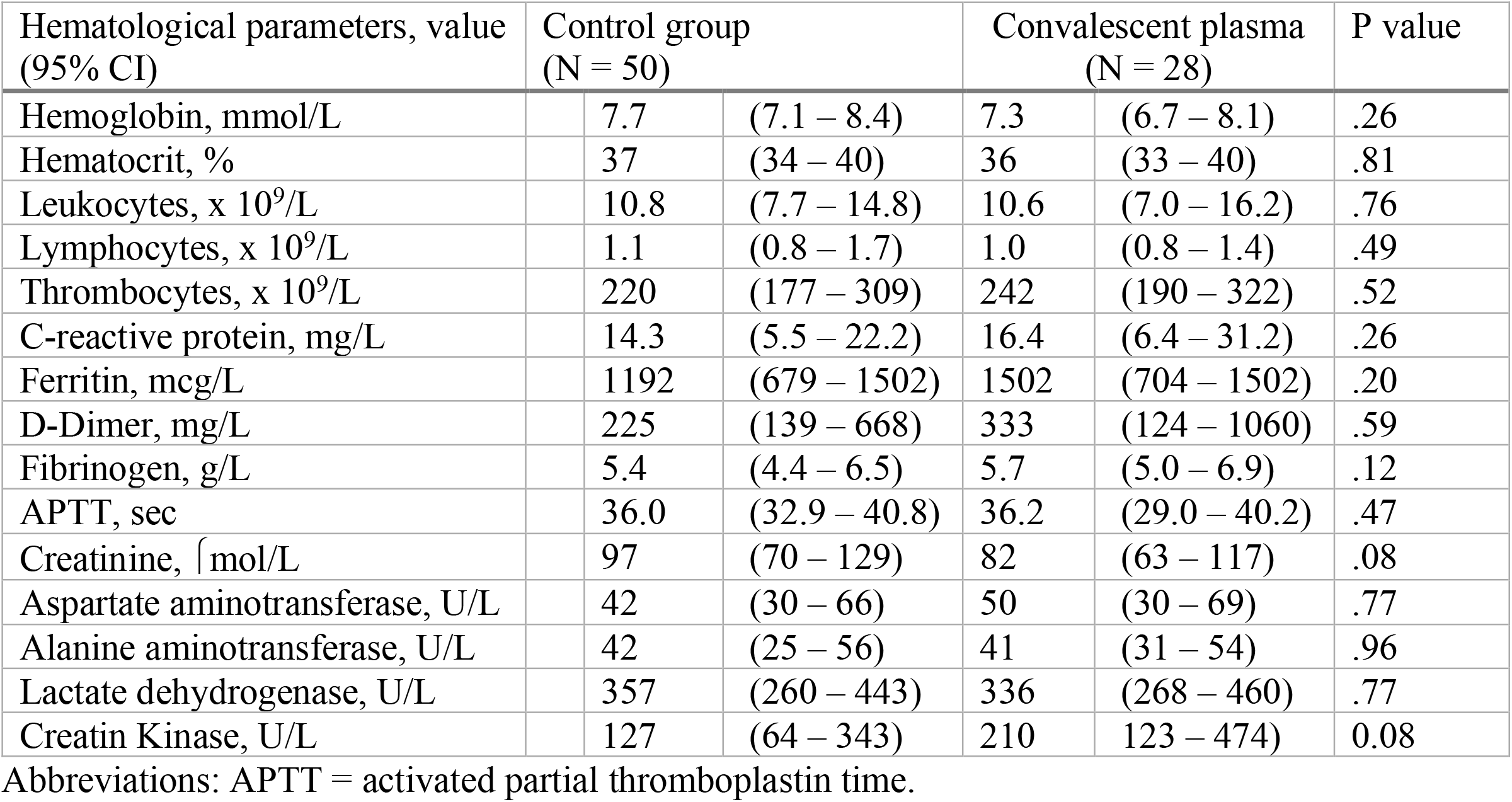
Comparison of Laboratory findings between the intervention (convalescent plasma) group and the control (dexamethasone) group at baseline.

### Convalescent plasma donors

All donors who were selected and screened according to the guidelines for CP donation tested positive on qualitative testing for Abs. The mean OD(± sd) was 2.66 (± 0.23). Based on studies available at the beginning of the trial, describing the relation between OD levels of the CP in relation to its virus neutralizing efficacy together with the rather early harvesting after illness of the donor, it was assumed to be effective CP ^13,17^.

### Primary outcome: 28-Day Mortality

Survival probability was significantly higher in the CP group compared to the control group with standard care (P=0.027) (Figure 3a). At day 28 on the ICU, mortality had occurred in five out of 28 (18%) of the intervention group versus eighteen out of fifty (36%) in the control group. When stratifying the therapy groups into disease severity as determined at baseline, the survival probability in the control group with life-threatening disease was significantly lower compared to the other three conditions, with a log-rank p-value of 0.0051 (Figure 3b). These results suggest a protective effect of CP therapy, which is most pronounced in the life-threatening group. Although there seems to be some effect of baseline disease severity on mortality probability, this was not statistically significant and CP therapy remained positively associated with reduction of mortality even when correcting for this. Univariate hazard ratios and 95% CI and log-rank p-value revealed that age, the presence of diabetes, and the severity of COVID-19 at baseline and treatment with CP had a significant impact on mortality (Table S1). When corrected for these four significantly influencing covariates, a higher survival probability was still found for the convalescent plasma intervention group, with a hazard ratio (HR) of 0.22 (95%CI, 0.074-0.067) (Figure 3c). Age was the only other covariate that increased the risk of mortality significantly (HR, 1.08 [95%CI, 1.022-1.14]; P = 0.006). The severe COVID-19 group had a non-significant trend towards reduced mortality compared to the life-threatening group with a HR of 2.39 [95% CI, 0.82-6.97]; P =0.11.

**Figure 3.**
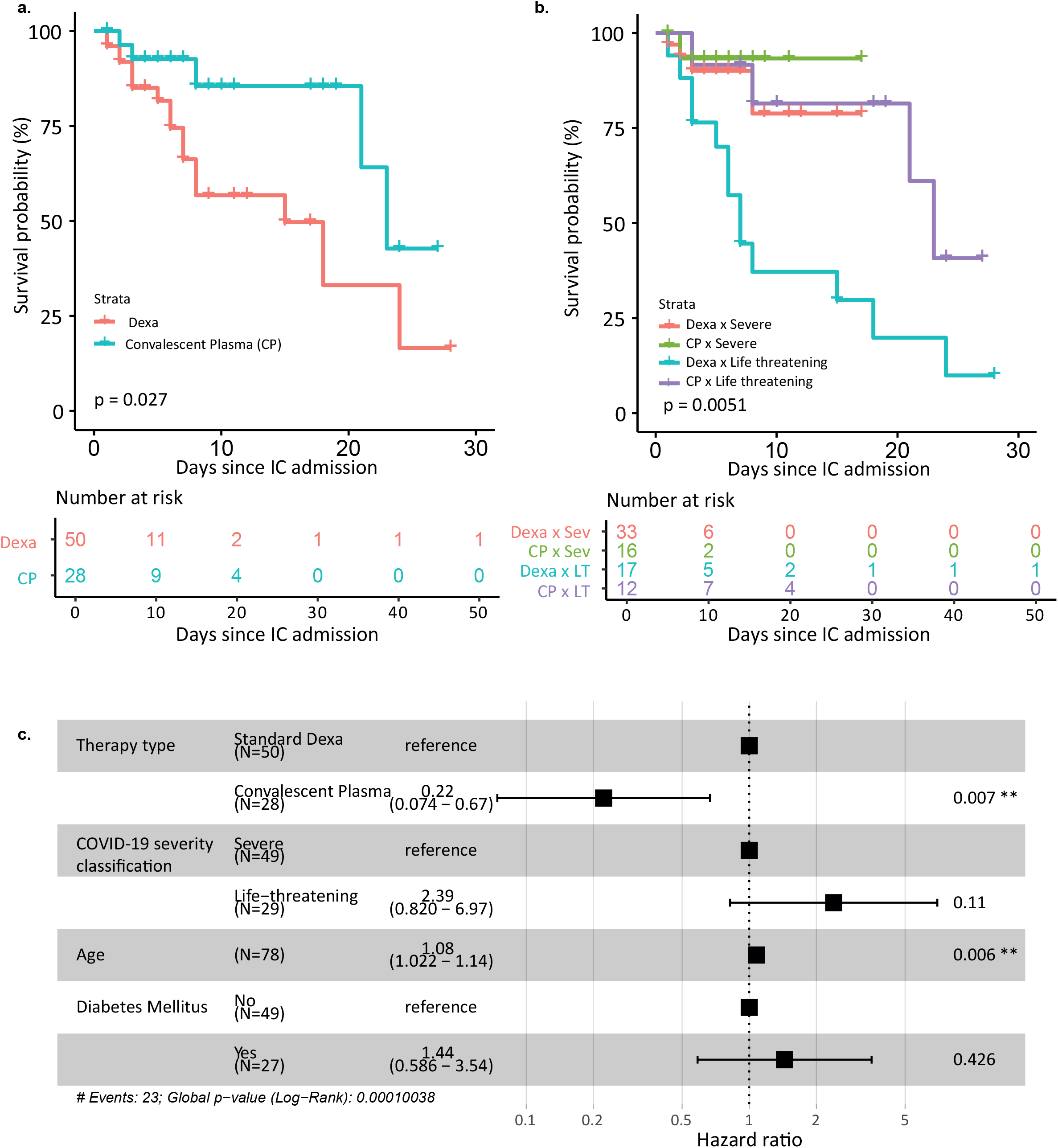
Survival probabilities during IC admission for convalescent plasma and standard dexa treatment group. **a)** Kaplan Meier curve of complete dexa (n=50) and convalescent plasma (n=28) treatment groups. Right-censoring took place when a person was dismissed from the IC before the last measured timepoint, death was counted as an event. The longest IC admission duration in the convalescent plasma group was 27 days, while it was 50 days in the standard dexa group. **b)** Kaplan Meier curve of both treatment groups, split into the COVID19 severity classification. Censoring took place when a person was dismissed from the IC before the last measured timepoint, death was counted as an event. The p-value of the log-rank signed test is reported directly in the figure. **c)** A multivariate cox proportional hazards analysis was performed with the univariately significant vairables. Hazard ratios and 95%CI and log-rank p-value are reported.

### Secondary outcome: CRX score and PF ratio

#### Chest radiographic findings

To identify early changes in clinical response, the CXR score was determined for each patient upon ICU admission (Day 0) and 48 hours after the treatment initiation (Day 2). For the control group, no changes were observed in CXR score between day 0 and 2 in the severe disease category (Day 0: 7.5 [4.8-11], Day 2: 8 [6-12], median change: 1 [-1 – 2], p=0.19), or in the life-threatening category (Day 0: 11 [3-12], Day 2: 8 [4-13], median change: 0 [-3 – 2], p=0.92) (Figure 4a). In the CP group, a downward trend was observed in both the severe (Day 0: 9 [6.3-10]; Day 2: 7 [4.8-8.3], median change: −3 [-4 – −1.5], P=0.07) and life-threatening (Day 0: 6 [5-6.8]; Day 2: 2.5 [2-3.8], median change: −3 [-4.8 – −1], P=0.07) categories. The effect size was determined for both groups by calculating the difference in CXR score between day 0 and day 2 (delta CXR) (Figure 4b). In the severe disease category, the delta CXR after CP treatment, i.e. median −3 points, was significantly greater than in the control group (median 1 point, P=0.0013). In the life-threatening disease group, no significant improvement in CXR score was found between the CP (median −3 points) and control group (median 0 points, P=0.48).

**Figure 4.**
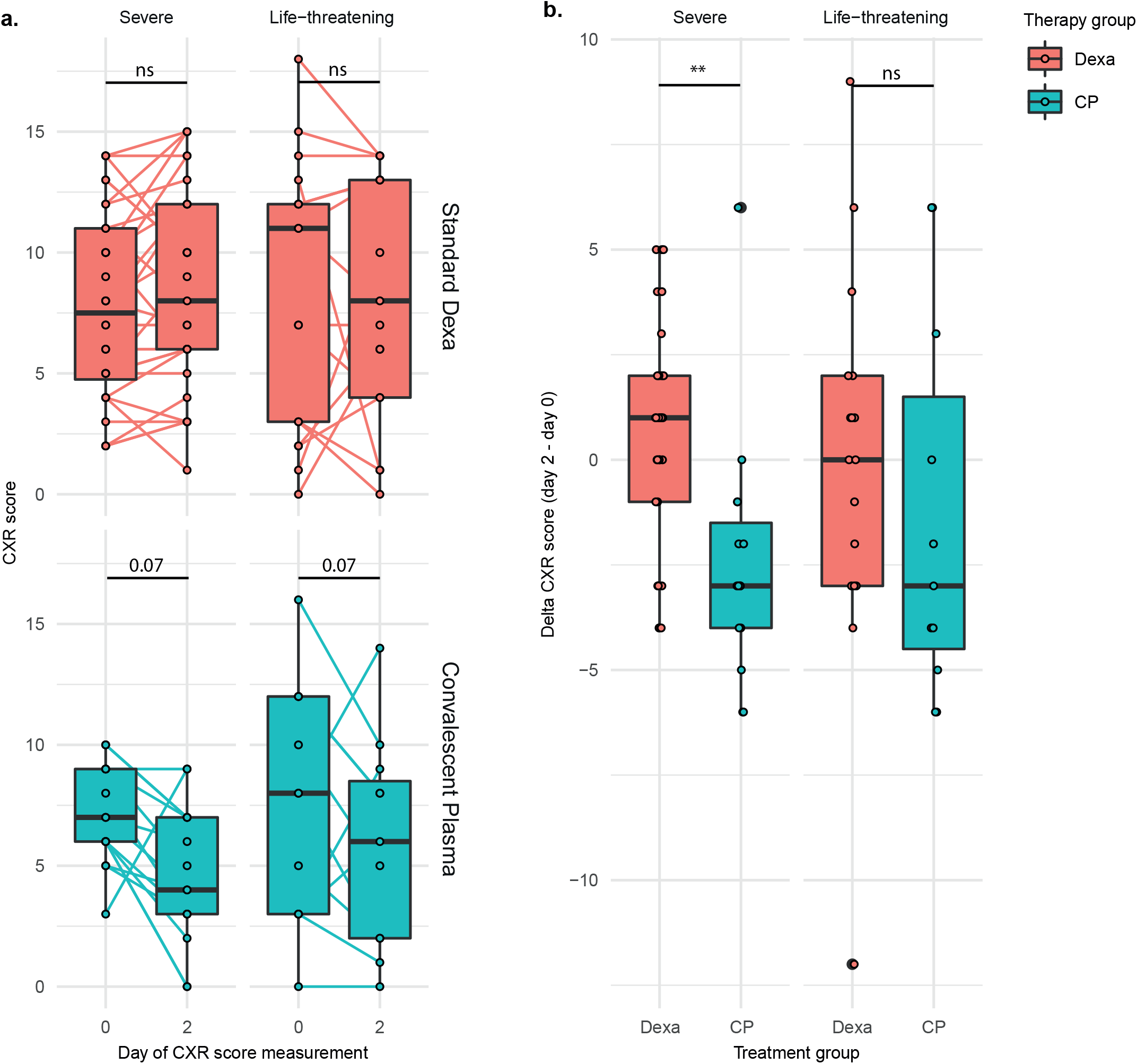
CXR score improvement in convalescent plasma and standard dexa treatment group. **a)** Change of CXR score from day of IC admission (day0) and one day after treatment start (day2). The standard dexa treatment (red) and convalescent plasma treatment (blue) groups are categorized into severe and life-threatening at the time of IC admission, based on the COVID19 severity classification described by Ling et al. Differences between the two time points were calculated using a two-sided paired t-test. **b)** The delta CXR score in the standard dexa and convalescent plasma group. The delta CXR score is calculated by subtracting the score of day 0 from the score on day 2. The differences between the two groups were calculated using an unpaired t-test with variance correction, * p <0.05, **p<0.01, *** p<0.001. p-values of borderline significant tests are reported directly in the figure. NS: not significant.

#### PF ratio

Besides changes in CXR score, we also evaluated the impact of CP on pulmonary oxygen exchange capacity (PF ratio). Significant improvement of PF ratio (PFR) after 48 hours was observed both in the control group (Day 0: 94.4 [75.4-126.5]; Day 2: 151.4 [100.3-231.8], median change: 35.3 [14.6-83.3], P<0.001) as well as the CP group (Day 0: 150.8 [93.1-300.5]; Day 2: 286.7 [201.7 – 390], median change: 100.25 [29.2-168], P=0.03) (Figure 5a). In the life-threatening category, no improvement was observed in the control group (Day 0: 99.5 [81.3-132]; Day 2: 99.4 [74.8 – 183.8], median change: 18.5 [-19.9 – 48.6], P=0.26). An upward trend was observed in the CP group (Day 0: 166.7 [97.0 – 236.1]; Day 2: 249 [189.2-397.5], median change:110.5 [-3.5 – 231], P=0.08), but this just failed to reach statistical significance.

**Figure 5.**
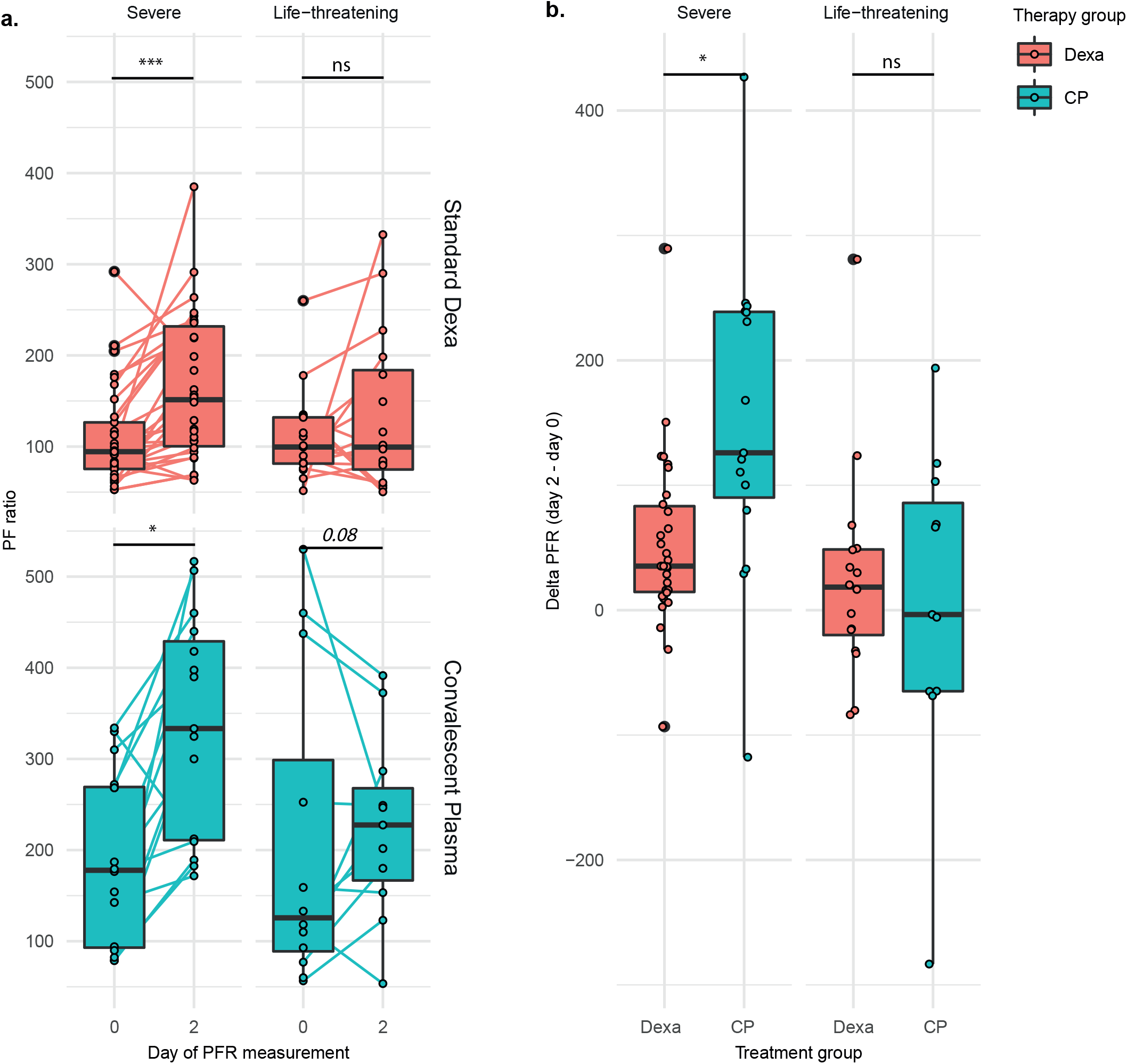
PF rate improvement in convalescent plasma and standard dexa treatment group. **a)** Change of PFR from day of IC admission (day0) and one day after treatment start (day2). The standard dexa treatment (red) and convalescent plasma treatment (blue) groups are categorized into severe and life-threatening at the time of IC admission, based on the COVID19 severity classification described by Ling et al. Differences between the two time points were calculated using a two-sided paired t-test. **b)** The delta PFR in the standard dexa and convalescent plasma group. The delta PFR is calculated by subtracting the score of day 0 from the score on day 2. The differences between the two groups were calculated using an unpaired t-test with variance correction, * p <0.05, **p<0.01, *** p<0.001. p-values of borderline significant tests are reported directly in the figure. NS: not significant.

A comparison of the PFR improvement after the standard (median 35.3) or CP treatment (median 151.6) revealed a significantly greater improvement in severe CP group (P=0.011) (Figure 5b). In the life-threatening group, no significant difference in PFR improvement was found between the control (median 18.5) and convalescent plasma group (median 166.7, P=0.63).

## Discussion

The COVID pandemic has had a major health and socio-economic impact on LMICs, including Suriname. Many potential treatment options being developed in high-income countries, including the use of CP, are not available in low-resource settings. Here, we report the interim results of a non-randomized intervention trial in Suriname using COVID-19 convalescent plasma obtained via gravity-filtration. The primary outcome showed a significantly higher ICU survival probability in the CP group compared to the control group, with a hazard ratio of 0.22. Significant improvement was also observed for the secondary endpoints, i.e. changes in PF ratio and CXR scores 48 hours after treatment over baseline, as markers for early clinical treatment effects.

Based on 32 earlier studies on outbreaks of SARS coronavirus infection and severe influenza, consistent evidence was found for beneficial effects of CP therapy on survival ^18^. The mortality reduction that we observed is in line with other reports on CP treatment in severe COVID-19 ICU patients. A review of nine studies showed that CP treatment significantly reduced mortality of ICU patients with COVID-19, which was more pronounced in severely ill patients than in critically ill patients. CP treatment in severe COVID-19 cases was concluded to result in clinical improvement with limited adverse events^19^. In a propensity score–matched control study of Mount Sinai, survival also improved in CP recipients, including improvement of oxygenation^20^. However, in a prospective open label randomized trial by Li performed in the Wuhan region in China, 28-day mortality was not significantly reduced (15.7% vs 24.0%; OR, 0.65; 95% CI, 0.29-1.46; P =.30)^21^. In that study, clinical improvement within 28 days was observed in the severe convalescent plasma group compared to the control group, but not in the subgroup with life-threatening disease (HR, 0.88; 95% CI, 0.30–2.63; P = 0.83). Since the interaction by disease severity failed to reach statistical significance; Li concluded that there was no benefit overall of CP treatment^6^. A recent randomized trial by Simonovich, no significant benefit of CP at 30 days with regards to clinical outcome, with 11% mortality observed both in the CP group and in the placebo group^22^. This is in contrast with our findings, where we observed a strong significant overall effect of CP, which remained significant in the life-threatening group although with an almost two fold increased risk on mortality when compared to the severe group.

Due to differences in healthcare, patient demographics, co-morbidities, circulating SARS-CoV-2 variants and many other factors it is difficult to directly compare results from other trials with our study. There may be several explanations for why we observed a strong treatment effect. A key factor might be the composition of CP that was used ^23,24^. The composition CP likely varies according to the method used to separate plasma from blood, i.e. by mechanical centrifugation or by filtration^25–27^. During centrifugation, plasma factors may be damaged or inactivated by shear forces generated by centrifugal or pressure forces ^28^. In contrast, the gravity-based filtration method we used to produce CP is based on low pressure and low shear forces. Moreover, the membrane pores are 3 to 10 times larger than those of conventional plasmapheresis membranes, which may affect large molecules differently^26^. During the acute phase of SARS-CoV-2 infection, patients start producing antibodies including IgA, IgM and IgG against viral epitopes. Whereas IgM antibodies wane rapidly from circulation, IgG becomes the dominant antibody isotype in blood ≥6 weeks after initial onset of disease ^21^. Additionally, blood from convalescent patients who were admitted to ICU because of severe or life-threatening COVID-19 contained almost 4-8 fold higher antibody concentrations than mild or moderate COVID-19 patients^23^. Since we isolated plasma from convalescent ICU patients with a median of 3 weeks after recovery by gravity-filtration, it is likely that CP from these donors contains higher concentrations of virus-neutralizing antibodies, including IgM, than CP from patients who recovered from less severe COVID-19 disease isolated at a later stage after recovery. It has previously been reported that patients that are severely affected with respiratory failure produce less IgM ^29,30^. At present not much is known about the effective dose of CP and whether repetitive doses are required or not. Mostly, CP dose is based on the practical supply of CP by blood banks as 220ml bags. In the current trial, two doses of 220ml were infused on one day, which is higher than the study by Li *et al*. where most patients received a single infusion of 200 ml of CP ^31^. The average age of patients in the CP and control group in our study was 53 and 54 years, respectively, which is much lower than in other studies where the mean age has been reported above 60 or even 70 years of age ^32,33^. Since increasing age was associated with reduced survival probability in our study, age should be taken account when interpreting study results of CP therapy. The effects of age may extend beyond CP treatment, since the cardiac surgery DECS trial indicated that dexamethasone may only be beneficial in treating respiratory failure when given to patients below 75 years of age^34–36^. A significant decrease in mortality was seen with the use of dexamethasone for up to 10 days in patients requiring respiratory support as described in the RECOVERY study^1^. The effect of prolonged immunomodulation could thus be an asset in the recovery from severe or life threatening COVID-19 disease. This effect was not observed in our control group, probably due to the already very high severity of COVID-19 upon admission to the ICU. The increase of other clinical parameters such as PF ratio and decrease of CXR score by resolving pulmonary oedema may represent first step in alleviating the severity of respiratory failure, thereby increasing survival probability.

CP effects on mortality reduction were most obvious in the life-threatening group, which was associated with early improvement in delta PF ratio and CXR score 48h after treatment initiation. Although it remains unclear which plasma factor or combination thereof may drive clinical improvement, it may be a direct effect of virus-neutralizing antibodies ^37^.

Alternatively, but not mutually exclusive, the rapid clinical improvement may also be due to direct immunomodulatory effects of anti-inflammatory plasma factors such as C1-esterase^38^. Restoring endothelial barrier function and improvement of alveolar-capillary oxygen transmission and oxygen diffusion capacity are possible treatment effects of CP ^39^.

Studies on novel therapeutics interventions in LMICs, including the use of CP, are generally faced with many practical barriers, both with regards to clinical and laboratory equipment. For instance, centrifugation-based plasmapheresis requires sophisticated and difficult-to-transport equipment. In the absence of such equipment, harvesting of CP in our study was made possible by a novel filtration device, i.e. the HemoClear device. Although initially indicated for blood salvage, the portable device proved an accessible method for plasma collection in low-resource settings as there is no need for electricity and the device can be used at the bedside. Both CXR and PF scoring may facilitate evaluation of efficacy of treatments such as CP and clinical decision making in a LMIC setting. Although we have not yet validated the use of chest x-ray as a surrogate method to evaluate the efficacy of CP, results from our study are in line with the recent validation study by Borghesi, et al, which showed that the CXR score was significantly higher in patients who died than those discharged from the hospital. This suggests that CXR scoring may represent a useful and practical tool to objectively score clinical improvement and identify early signs of improvement^40^.

This study is limited by the non-randomised design, which may have resulted in differences between the CP and the control group. As CP was not always readily available during the second epidemic, selection bias could be present. Furthermore, CP therapy was initiated three days after ICU admittance. As such, treatment bias may therefore exist since the CP group consisted of more life-threatening patients than the control group. However, it should be noted that this bias would be expected to decrease any beneficial effect of CP treatment. Since we observed an opposite effect, one could argue that that CP was highly efficacious. Another limitation is the relative small number of patients. Based on the results of this trial, new patients are being enrolled which may provide further evidence on the potential beneficial effects of CP. Further characterization of the CP used in this study is needed to fully ascertain its efficacy and correlate variation in plasma factors to clinical outcome in CP recipients.

In summary, COVID-19 remains a global health threat and reliable treatment is crucial for reducing mortality and the burden on global health care. Access to CP therapy in a low-resource setting was enabled by the novel filtration device Hemoclear, which was easy to implement in an ICU setting and was used without adverse effects on both the donor as the CP recipients ^41,42^. Equitable access to such methods allows readiness in case of viral mutations or new pandemics. The use of simple and available methods such as chest x-ray and calculated PF ratios allowed early assessment of treatment effects. SARS-CoV-2-specific therapies, including CP from recovered patients, could be highly effective options to treat COVID-19 in the absence of widespread vaccination. As such, CP therapy may help bridge the gap until sufficient vaccination coverage has been reached.

## Supporting information

COVID-19 Convalescent Plasma

## Data Availability

The study data are available upon request.

## Acknowledgements

We would like to thank Dion Osemwengie for her support in training on use of the blood filter and contributions to the plasma acquisition protocol.

## Data availability

The data used to support the findings of this study are available from the corresponding author upon request.

---

**Table S1:** univariate hazard ratios of patient specific variables on death during IC admission. A univariate cox proportional hazards analysis was performed for each variable. Hazard ratio, 95% confidence interval and p-value are described. Bolded variables are significantly associated with death and included in the multivariate cox proportional hazards analysis, variables in italics are borderline non-significant and not included in further analysis. The COVID-19 severity classification is based on the classification described by Ling et al.*measured at time of IC admission

|  | *HR* | *95%CI* | *p-value* |
| --- | --- | --- | --- |
| **Age** | **1.1** | **(1-1.1)** | **0.0025** |
| BMI | 0.99 | (0.92-1.1) | 0.66 |
| Sex | 0.66 | (0.25-1.7) | 0.4 |
| **COVID-19 severity classification** | **3** | **(1-8.4)** | **0.042** |
| Treatment start day | 0.94 | (0.87-1) | 0.12 |
| **Therapy type** | **0.34** | **(0.12-0.92)** | **0.035** |
| D'dimers* | 1 | (1-1) | 0.26 |
| DeltaCXR* | 1 | (0.9-1.1) | 0.88 |
| DeltaPFR* | 1 | (0.99-1) | 0.38 |
| SAP* | 0.99 | (0.97-1) | 0.46 |
| DAP* | 0.99 | (0.95-1) | 0.54 |
| CRP* | 0.98 | (0.95-1) | 0.38 |
| Ferritin* | 1 | (1-1) | 0.29 |
| Fibrinogen* | 1 | (0.96-1) | 0.94 |
| Hb* | 0.83 | (0.58-1.2) | 0.31 |
| *Creatin\** | *1* | *(1-1)* | *0.065* |
| LDH* | 1 | (1-1) | 0.13 |
| Thrombos* | 1 | (1-1) | 0.96 |
| *Hypertension* | *2.3* | *(0.94-5.7)* | *0.069* |
| Ischemic heart disease | 0.96 | (0.22-4.2) | 0.95 |
| **Diabets Mellitus** | **2.4** | **(1-5.5)** | **0.047** |
| Kidney Failure | 1.2 | (0.4-3.5) | 0.77 |
| Stroke | 2.1 | (0.47-9.7) | 0.33 |

**Video S1:**
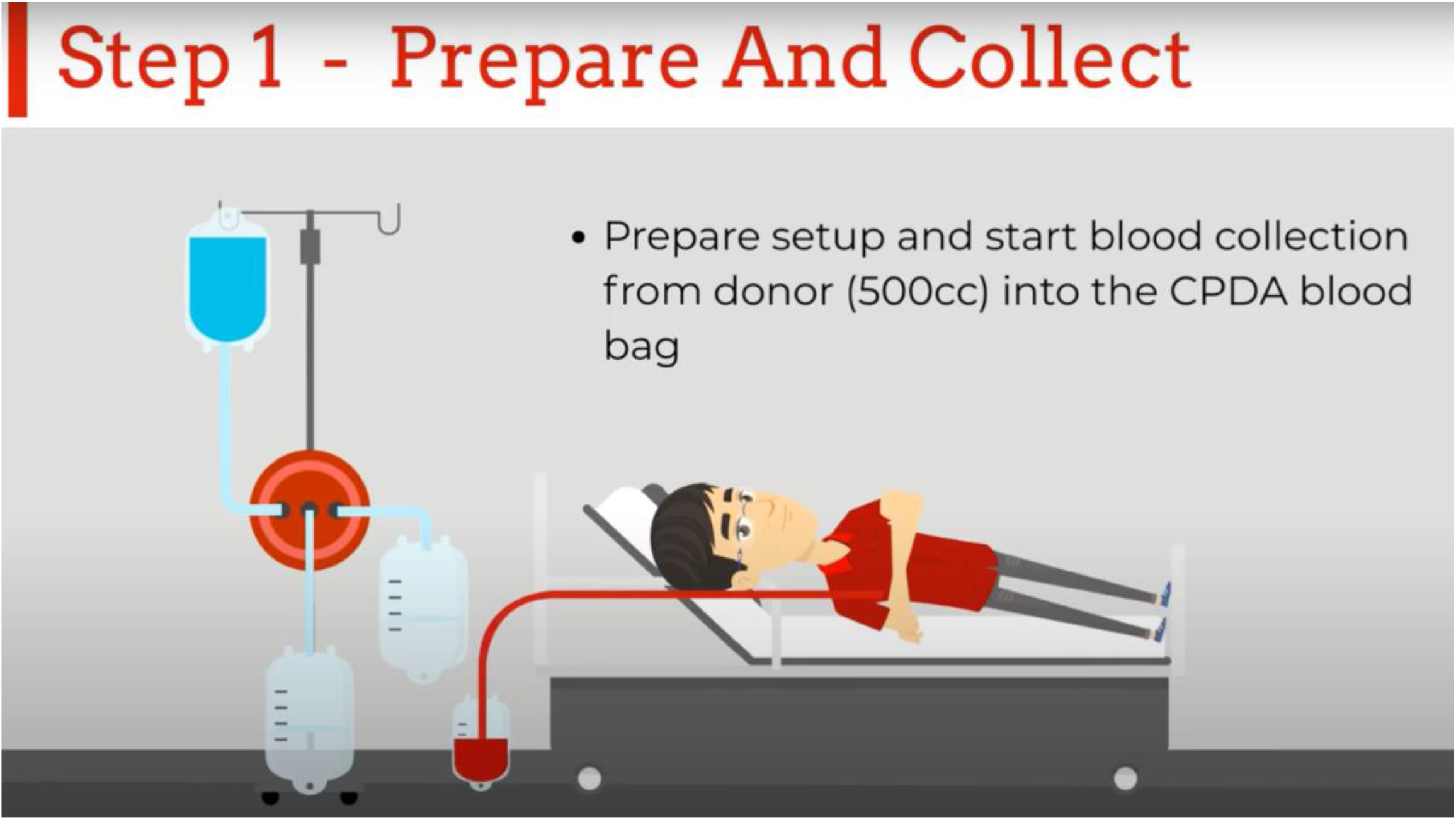
Infographic on the plasma collection method. This video can also be found online at: https://www.youtube.com/watch?v=gQGh-PtfBLk

